# Mapping the Psychopathic Brain: Divergent Neuroimaging Findings converge onto a Common Brain Network

**DOI:** 10.1101/2024.09.12.24313535

**Authors:** Jules R. Dugré, Stéphane A. De Brito

**Affiliations:** School of Psychology and Centre for Human Brain Health, University of Birmingham, Birmingham, UK; Institute for Mental Health, Centre for Developmental Science, Birmingham Centre for Neurogenetics, University of Birmingham, Birmingham, UK

**Keywords:** Psychopathy, Neuroimaging, Meta-analysis, Brain Network, Connectivity

## Abstract

Psychopathy is a personality disorder characterized by a constellation of interpersonal, affective, lifestyle, and antisocial features. However, its neural underpinnings remain poorly understood because functional neuroimaging studies have produced disparate findings. Here, we tackled this lack of replication by investigating whether peak coordinates of studies on psychopathy could in fact map onto a common functional connectivity network. An updated meta-analysis of 23 functional neuroimaging studies (534 cases vs 594 controls) first revealed no significant regional spatial convergence. However, using functional connectomes of 1,000 healthy participants, we demonstrated that the heterogeneous study findings do indeed converge onto a common brain network with a replicability reaching up to 85.2% across studies. We subsequently showed strong associations between this Psychopathy Network and a lesion network of 17 lesion sites causally linked to antisocial behaviors, as well as its association with neurotransmission systems and genetic markers previously implicated in the pathophysiology of psychopathy. Taken together, our study highlights the importance of examining the neural correlates of psychopathy from a network perspective, which can be validated using a multilevel approach, encompassing neural, genetic and neurochemical data. Ultimately, this approach may pave the way for novel and more personalised treatments.

## Main

Psychopathy is a personality disorder characterized by a constellation of interpersonal (e.g., grandiose, arrogant, deceitful and manipulative), affective (e.g., superficial charm, shallow affect, callousness, lack of empathy and guilt), lifestyle and antisocial symptoms (e.g., need for stimulation, irresponsibility, criminal versatility, early behavioral problems) ^1^. It is estimated that around 1% of the general population meets the diagnostic criteria for psychopathy, but its prevalence rate reaches up to 25% in prisons ^1^. Psychopathy is associated with a wide range of negative psychosocial and behavioral outcomes, including high rates of criminal reoffending, which impose a substantial financial burden on society each year ^2^. To better understand its development, researchers have formulated various theoretical models, encompassing emotion-based ^3–5^, cognitive-based ^6,7^, and neural-based ^8,9,10^ perspectives. While psychological and criminological correlates of psychopathy have been well described over the past decades, investigating the neurobiological underpinnings of psychopathy may offer further insights into its pathogenesis.

Recent work has advanced our understanding of the genetic and neurobiological correlates of psychopathy. Behavioral genetic studies have shown the heritability of its development ^11^while molecular genetics and neurochemical research has identified potential genetic markers (e.g., MAOA, BDNF, COMT, HTR1B, HTR1A, OXTR, SLC6A4) and neurotransmission systems (e.g., dopamine, serotonin, norepinephrine) implicated in the disorder^12–14^. By contrast, despite the considerable attention functional neuroimaging has received to understand the brain (dys)functions of psychopathy, the neural substrates of the disorder remain poorly understood.

Functional magnetic resonance imaging (fMRI) work has provided support for alterations in brain functioning during emotional responsiveness, reinforcement-based decision-making, and attentional tasks in psychopathy ^1^. Indeed, two recent meta-analyses of fMRI studies have identified alterations in brain responses in psychopathy of the amygdala, dorsomedial and lateral prefrontal cortex, anterior insula (aINS), and anterior and posterior cingulate cortices (ACC/PCC) ^15,16^. Importantly, lesion studies in humans have also shown that damage to these brain regions (including the ventromedial prefrontal cortex [vmPFC]), may cause a clinical profile closely resembling psychopathy (e.g., blunted affect, lack of empathy, impulsive behaviors) ^17–20^. This personality disturbance, initially termed as “*pseudopsychopathy*” ^21^ or “*acquired sociopathy*” ^20^, is associated with similar impairments to those seen in psychopathy such as reduced autonomic responses to social stimuli ^10^, impaired decision making ^22,23^ and reduced prosocial motivation ^24^.

While these regions appear to be potential candidate neural substrates for psychopathy, fMRI studies have been plagued by low replicability of these findings ^15,16^. This state of affairs has cast doubts on the robustness of the so-called neurobiological markers of psychopathy, prompting some researchers to state that: “*no reproducible evidence suggests that psychopathy is associated with a functional neurobiological profile”* ^25^ (p.6). Arguably, it is possible that variations in psychopathy symptoms, incorporating interpersonal, affective, lifestyle and antisocial features, may partly account for the heterogenous findings across studies^26^. Crucially, however, the poor replicability of fMRI findings is not limited to research on psychopathy but is common across the field of psychiatric neuroimaging. Addressing the replicability crisis in neuroimaging is imperative to advance our understanding of the neurobiological underpinnings of psychopathy.

One promising approach to circumvent the weak overlap at a regional level is to consider the brain as having a modular organization of densely interconnected regions ^27,28^. Indeed, cumulative evidence suggests that variability of the blood oxygen level-dependent (BOLD) signal in different brain regions may reflect their strength of functional connectivity across the brain ^29–32^. That is, brain regions would require more energy (e.g., supplies in oxygen, glucose) if they show greater functional connectivity with the rest of the brain ^33,34^. Averaging BOLD signals across task conditions and subjects may thus reduce the importance of non-hubs regions (those with fewer connections), which may nevertheless be essential to our understanding of mental functions and psychopathologies ^35^. Researchers have thus recently adopted a more distributed view of brain functioning by using *normative functional connectivity* to map heterogeneous brain regions of the same phenotype onto a common network ^36^. This approach has been successfully applied to heterogeneous brain lesions linked to criminal offending ^37^ as well as to heterogenous findings reported across studies on substance use disorder ^38^. Investigating neurobiological markers of psychopathologies through a distributed view of brain functioning has the potential to enhance the translational value of fMRI data by, for example, identifying novel targets for neuromodulation and psychopharmacological treatments ^39^.

Here, we first conducted an updated meta-analysis of fMRI studies on psychopathy which revealed no significant regional convergence across studies. We thus investigated for the first time whether the heterogeneity of brain regions reported across studies may in fact converge onto a common brain network. Leveraging existing large-scale fMRI data, combined with state-of-the-art developments in normative modeling, we provide the first evidence for a Psychopathy Network. As an indicator of convergent validity, we demonstrate its strong associations with a lesion network causally linked to antisocial behaviors^37^, along with neurotransmission systems and genetic markers previously implicated in the pathophysiology of psychopathy.

## Results

### Activation Likelihood Estimation Meta-analysis

A total of 23 fMRI studies published were included in the current meta-analysis which comprised 534 cases versus 593 controls (see Table 1). Mean age across samples was 32.83 years (SD=6.87, range 19.5-44.6). From these studies, 11 reported recruiting participants through correctional/probation settings, 7 through forensic settings and 5 through community. First, coordinate-based meta-analysis using Activation Likelihood Estimation (ALE) on peak coordinates of the 23 studies (399 foci) revealed no significant spatial convergence (see Figure 1A). More thorough investigation, in which we modeled each peak coordinate with a 4mm sphere, revealed that the rate of *regional* spatial overlap between studies was very poor (i.e., maximum peak overlap: two studies out of 23, ≈ 8.70%) (Figure 1C).

**Figure 1.**
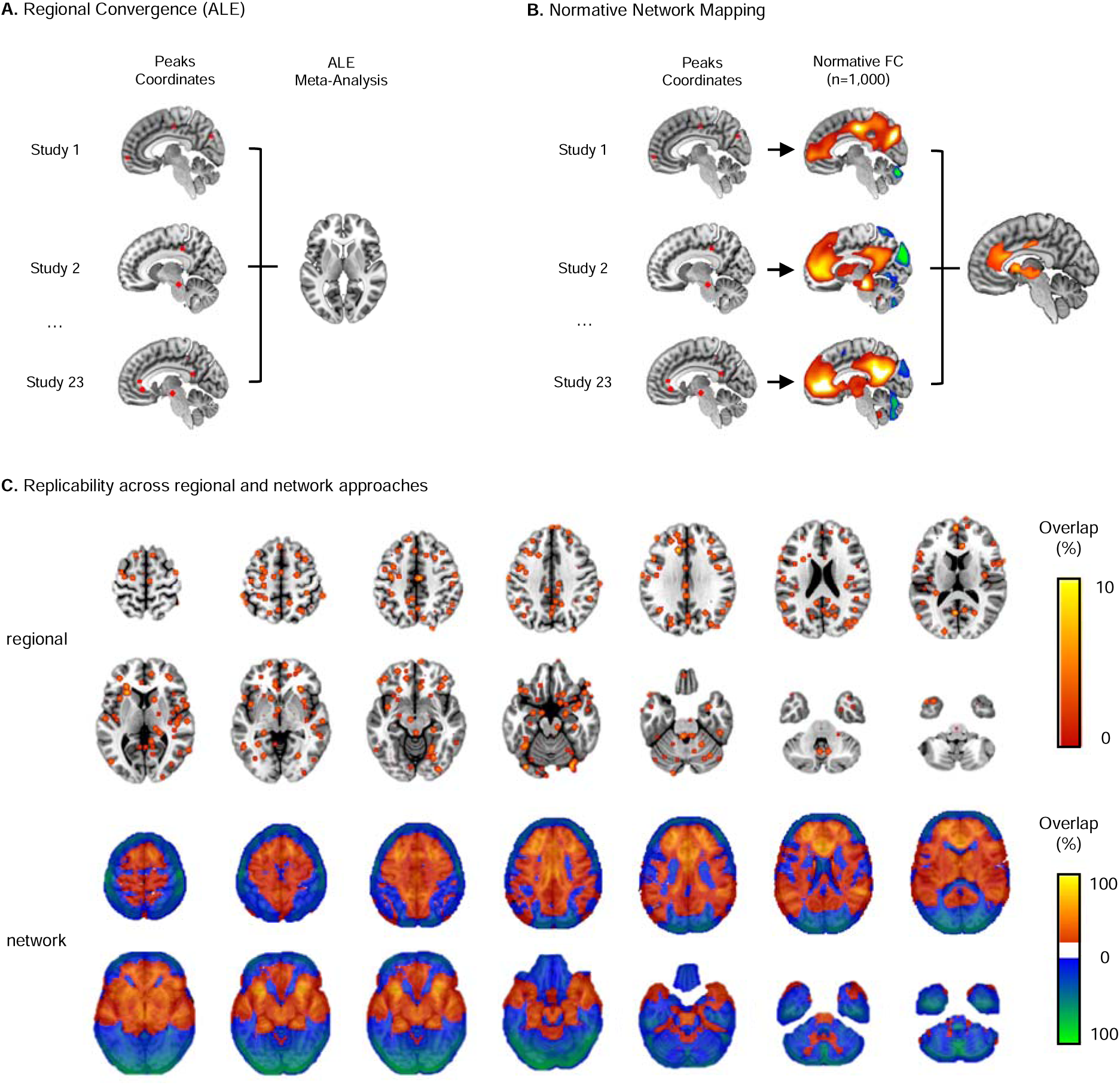
Spatial Convergence across neuroimaging studies on Psychopathy. **A.** Activation Likelihood Estimation meta-analysis was conducted on peak coordinates of 23 fMRI studies on Psychopathy which revealed no significant spatial convergence. **B.** Network mapping approach was conducted to identify the normative functional connectivity profile characterizing the 23 task-based fMRI studies. **C.** Displays overlap between binarized map of peak coordinates across studies (4mm sphere) resulting in low *regional* replicability (up to 8.70%)(upper row), and between binarized normative connectivity map across studies (t>5), resulting in high *network* replicability (up to 86.96% of studies)(lower row).

**Table 1.** Characteristics of the included fMRI Case-Control studies on Psychopathy (k=23)

| First Author, Year | Controls<br>(n=) | Sample Characteristics |  |  | Psychopathy Assessment |  | Neuroimaging Task |
| --- | --- | --- | --- | --- | --- | --- | --- |
|  |  | Cases<br>(n=) | Age | Setting | Measures | Total Score |  |
| Contreras-Rodriguez, 2014 <sup>85</sup> | 22 | 22 | 39.8 | Correctional | PCL-R | 27.8 | Facial Emotion Processing |
| Decety, 2015 <sup>86</sup> | 50 | 38 | 32.4 | Correctional | PCL-R | 30.0 | Interpersonal Scenarios |
| Deeley, 2006 <sup>87</sup> | 9 | 6 | 36 | Forensic | PCL-R | 29.3 | Facial Emotion Processing |
| Gregory, 2015 <sup>88</sup> | 18 | 12 | 40.1 | Correctional | PCL-R | 28.2 | Response-reversal task |
| Han, 2012 <sup>89</sup> | 16 | 16 | 24.4 | Community | PPI-R | 79.2 | Facial Emotion Processing |
| Harenski, 2010 <sup>90</sup> | 16 | 16 | 33.3 | Correctional | PCL-R | 31.8 | Morality Processing |
| Kiehl, 2004 <sup>91</sup> | 8 | 8 | 33.9 | Correctional | PCL-R | 32.8 | Lexical decision task |
| Larson, 2013 <sup>92</sup> | 25 | 24 | 33.1 | Correctional | PCL-R | 31.3 | Instructed Fear Task |
| Marsh, 2014<sup>A</sup> <sup>93</sup> | NA | NA | >18 | Community | PPI-R | 337.3 | Emotionally Statements |
| Meffert, 2013 <sup>57</sup> | 26 | 18 | 39.2 | Forensic | PCL-R | 32.3 | Emotional hand interactions |
| Mier, 2014 <sup>94</sup> | 18 | 11 | 44.7 | Forensic | PCL-R | 26.7 | Social Cognition Tasks |
| Müller, 2003 <sup>95</sup> | 6 | 6 | 33 | Forensic | PCL-R | 36.8 | Emotional Stimuli (IAPS) |
| Müller, 2008 <sup>96</sup> | 12 | 10 | 33.1 | Forensic | PCL-R | 30.5 | IAPS, Simon Paradigm |
| Pujol, 2012 <sup>97</sup> | 22 | 22 | 39.8 | Correctional | PCL-R | 27.8 | Moral Dilemma Task |
| Schulz, 2016 <sup>98</sup> | 19 | 31 | 31.6 | Correctional | PCL-R | 32.1 | Pavlovian fear conditioning |
| Sommer, 2010 <sup>99</sup> | 14 | 14 | 31.4 | Forensic | PCL-R | 28.6 | Emotion Attribution |
| Shane, 2018 <sup>55</sup> | 23 | 15 | 33.5 | Correctional | PCL-R | 31.9 | Emotional Regulation |
| Shao, 2017 <sup>100</sup> | 23 | 29 | 20.4 | Community | PPI-R | 336.9 | Face Familiarity (Lying) |
| Sethi, 2018 (A) <sup>101</sup> | 82 | 50 | 19.8 | Community | SRP-SF | - | Emotional face processing |
| Sethi, 2018 (B) <sup>101</sup> | 82 | 100 | 19.5 | Community | SRP-SF | - | Emotional face processing |
| Yoder, 2015 <sup>102</sup> | 34 | 28 | 32.6 | Correctional | PCL-R | 31.9 | Moral Evaluation |
| Volman, 2016 <sup>103</sup> | 19 | 15 | 37.8 | Forensic | PCL-R | 30.4 | Approach Avoidance |
| Deming, 2020 <sup>104</sup> | 33 | 26 | >18 | Correctional | PCL-R | >30 | Affective Perspective Taking |

Note. <sup>A</sup> Median split, without reporting number of participants per group.

### Normative Network Mapping

Using normative network mapping, which aims to examine convergence of brain regions through their functional brain connectivity networks, we found substantial overlap between studies. Indeed, overlapping study-level binarized maps increased replicability up to 86.96% (20 out of 23 studies) (Figure 1C, Table 2). This Psychopathy Network consisted of many key connectivity hubs including the perigenual anterior cingulate cortex (extending from the vmPFC to the posterior dACC area 24‘), subcortical regions (i.e., caudate nucleus, putamen, thalamus), midbrain (i.e., ventral tegmental area, red nucleus, raphe nuclei), bilateral fronto-insular cortices (ventral anterior insula, claustrum), bilateral superior frontal gyri (spanning the dmPFC, dlPFC and pre-supplementary motor area), posterior midcingulate cortex (pMCC, areas p24, 23c-d), and cerebellar lobules VIIIa-b and IX (TFCE FWE p<0.05)(Figure 1B, Table 2, see also Supplementary Figure 1 & Supplementary Table 1 for complete findings). These clusters showed moderate-to-high replicability when examining the average of voxels within cluster (up to 65.37%) and maximum overlap within cluster (up to 86.96%).

**Table 2.**
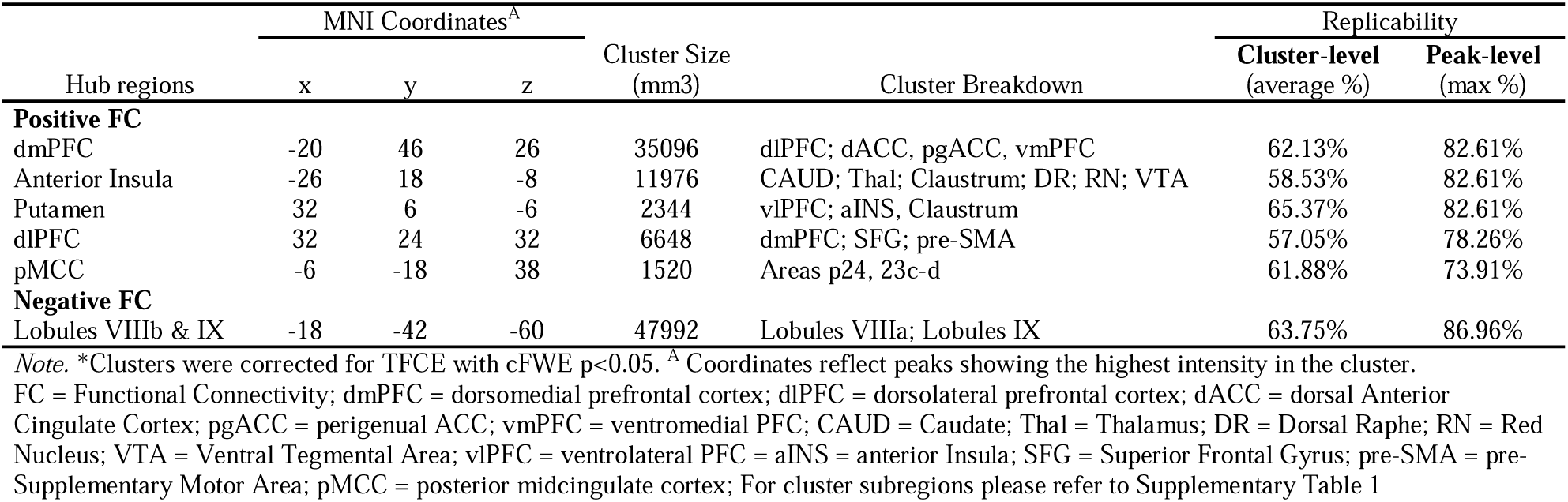
Functional Connectivity Hubs of Psychopathy Network and Replicability across studies.

### Contribution of Intrinsic Functional Connectivity Networks

We investigated whether the Psychopathy Network may be driven by specific intrinsic functional connectivity networks. Effect sizes (Cohen’s d) of Schaefer-400 parcels 7 Networks ^40^ and a subcortical network ^41^ were calculated as a measure of their contribution relative to the rest of the brain. We observed greater effect sizes within the default mode (Cohen’s *d* = 1.02), subcortex (Cohen’s *d* = 1.01), followed by ventral attention (Cohen’s *d* = .61) and fronto-parietal (Cohen’s *d* = .45) networks (Figure 2A).

**Figure 2.**
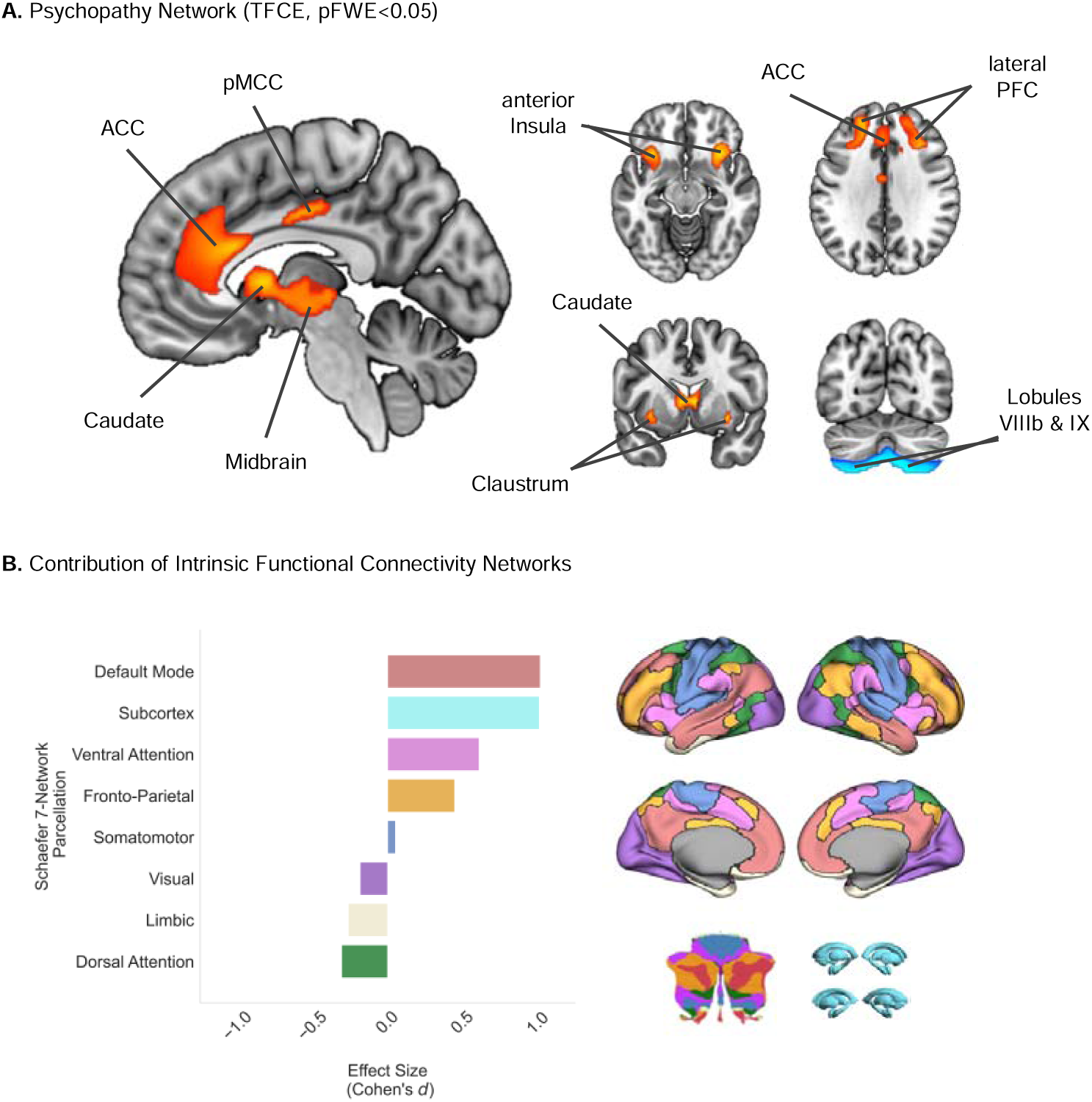
**A.** The Psychopathy Network (pFWE<0.05) included the following key hubs: the dorsal anterior cingulate cortex, midcingulate cortex, caudate nucleus, anterior insula, claustrum, lateral prefrontal cortex, thalamus, midbrain and the cerebellum (decreased connectivity). **B.** Investigating the contribution of 8 intrinsic connectivity networks highlighted the Default Mode Network (DMN) and Subcortex as core neural correlates the Psychopathy Network. The bar plot represents the effect size (Cohen’s d) of each network’s contribution. ACC = Anterior Cingulate Cortex; pMCC = posterior Midcingulate Cortex; PFC = Prefrontal Cortex.

### Functional Characterization

#### Brain Lesions Causally Linked to Antisocial Behaviors

We additionally examined the extent to which our findings map onto the network of brain lesions causally associated to antisocial behaviors, as identified by Darby and colleagues ^37^. We found a moderate-to-strong spatial correlation between both maps (r=.50, p<0.001 uncorrected). Of note, the spatial correlation increased to r=.61 (Figure 3) when the analysis was restricted to studies involving participants from correctional and forensic settings (i.e., individuals who have committed criminal offenses). This suggests notable neurobiological commonalities between the neural correlates of offenders with psychopathy and lesions resulting in antisocial behaviors (such as the DMN and subcortex). This provides neurobiological insights into how brain lesions may lead to clinical syndromes labeled *pseudopsychopathy* or *acquired sociopathy*.

**Figure 3.**
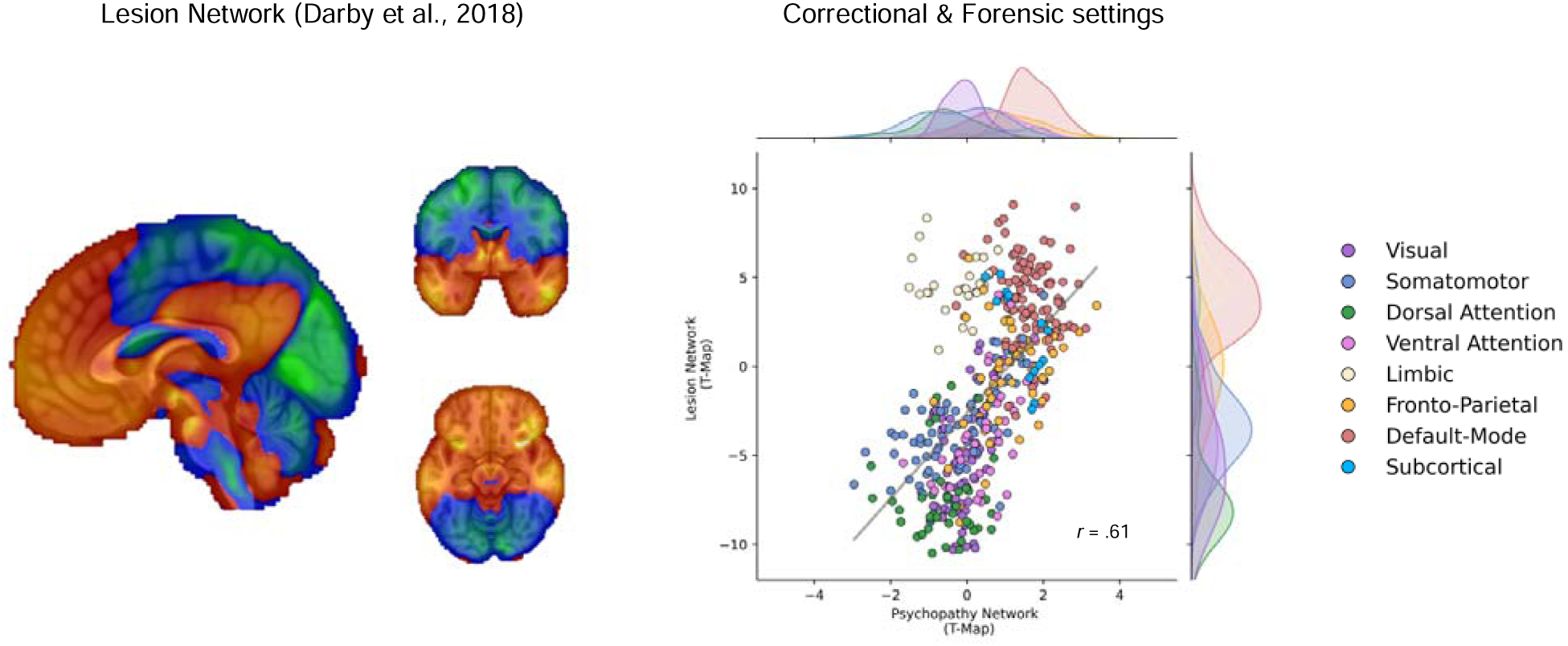
Association between the Psychopathy Network and Lesion Network underlying Antisocial behaviors, as reported by Darby and colleagues ^37^. A strong spatial association was found between our Psychopathy Network and the lesion network (*r*=0.50, p<0.001, not shown), particularly in studies including participants recruited from correctional and forensic settings (r=0.61, p<0.001). Common overlap between both maps was found primarily in the Default Mode Network (DMN) and Subcortex.

#### Mental functions and Neurotransmission systems

Spatial correlation was then conducted to investigate the association between the Psychopathy Network and mental functions^42^, and neurotransmission systems^43,44^ (Figure 4A). The Psychopathy Network showed strong associations with brain co-activity patterns subserving socio-affective processes including Social Inference (z’=.56, pFDR<0.05), Value-Based Decision-Making (z’=.56, pFDR<0.05), Social Representation (z’=.46, pFDR<0.05), and Motivation (z’=.46, pFDR<0.05) (Figure 4A and Supplementary Table 2). Spatial association with neurotransmission density maps revealed that the Psychopathy Network mainly correlated with dopamine receptor D_1_ (z’=.61, pFDR<0.05), serotonin transporter 5-HTT (z’=.60, pFDR<0.05), glutamate receptor mGLUR_5_ (z’=.52, pFDR<0.05), and dopamine transporter DAT (z’=.52, pFDR<0.05) (Figure 4A and Supplementary Table 2).

**Figure 4.**
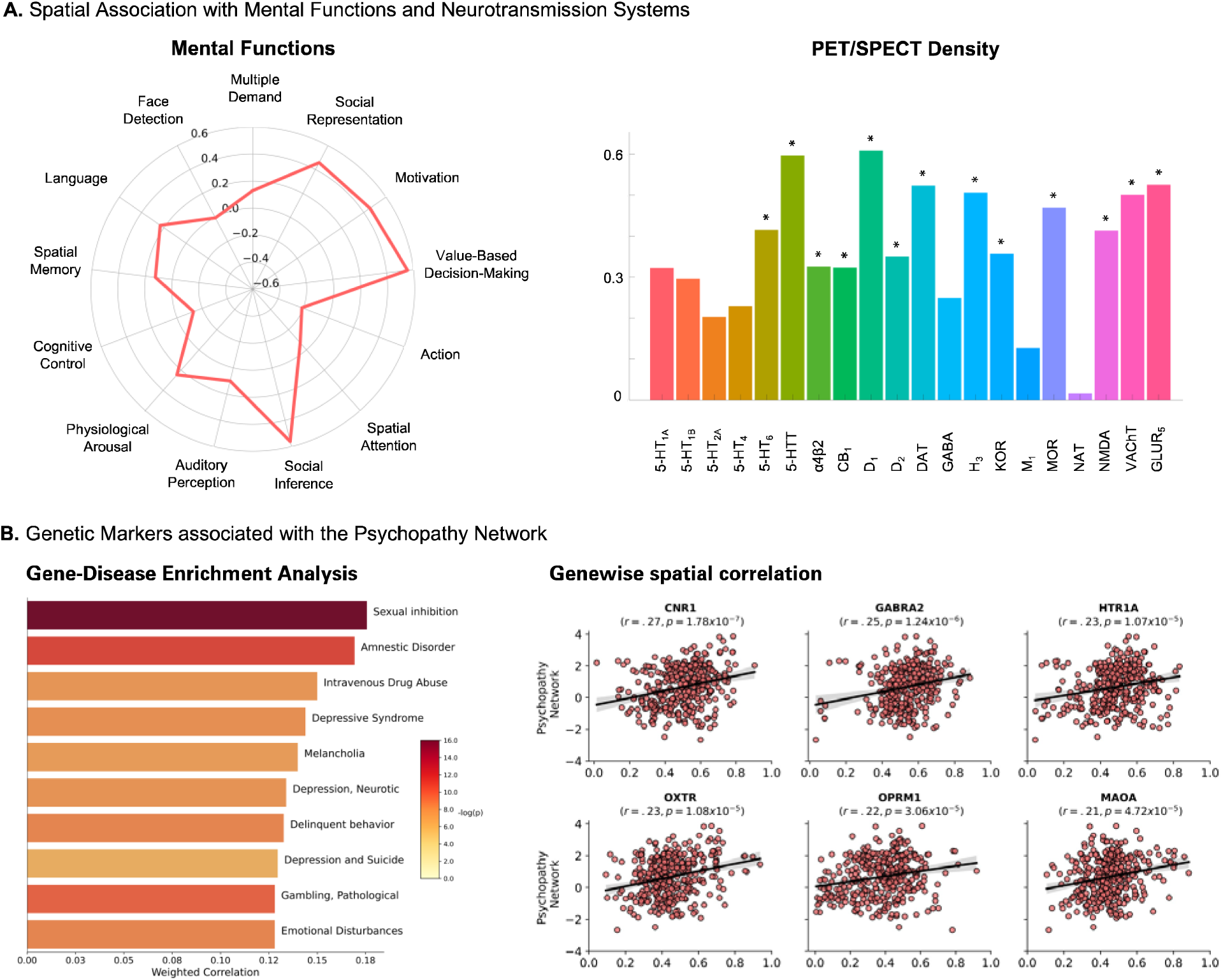
Functional Characterization of the Psychopathy Network. **A.** Psychopathy showed strong spatial associations with Social Inference (z’=.56) and Value-Based Decision-Making (z’=.56) derived from the brain-behavior ontology of task-based fMRI studies ^42^ as well as D_1_ (z’=.61), 5-HTT (z’=.60), mGluR_5_ (z’=.52), and DAT (z’=.52) density maps ^43,44^. Asterisks represent pFDR<0.05 correction. **B.** Gene-Category Enrichment Analyses identified 98 disorders strongly associated with Psychopathy Network. The left panel shows the top 10 disorders ranked by weighted correlations (i.e., log(number of genes+1). The right panel displays gene-wise spatial correlation patterns of the most strongly correlated genes identified across the 98 disorders.

#### Genetic Markers

Gene-Category Enrichment Analyses across the DisGeNET database^45^ revealed that the Psychopathy Network was significantly associated with genetic markers of 98 disorders (p<0.001, 5000 permutations) from which the top associated disorders were sexual inhibition (rho_weighted_=.18, p=1.11×10^-16^), amnestic disorder (rho_weighted_=.17, p=8.77×10^-12^), intravenous drug abuse (rho_weighted_=.15, p=2.04×10^-6^), depressive syndrome (rho_weighted_=.14, p=8.21×10^-9^), melancholia (rho_weighted_=.14, p=4.75×10^-8^), depression, neurotic (rho_weighted_=.13, p=1.41×10^-8^) and delinquent behavior (rho_weighted_=.13, p=3.35×10^-9^)(Figure 4B, and Supplementary Figure 2 & Table 4 for complete list of disorders).

Across those disorders, the most commonly occurring genes included BDNF (40.82%), COMT (31.63%), DRD2 (29.59%), MAOA (26.53%), APOE (21.43%), and SLC6A3 (21.43%) (Supplementary Table 3). Gene-wise spatial correlation patterns with the Psychopathy Network showed the strongest correlation with gene expression pattern of the CNR1 (z’=.27, p=1.78×10^-7^), GABRA2 (z’=.25, p=1.24×10^-6^), HTR1A (z’=.23, p=1.07×10^-6^), OXTR (z’=.23, p=1.08×10^-5^), OPRM1 (z’=.22, p=3.06×10^-5^), MAOA (z’=.21, p=4.72×10^-5^)(Figure 4B, Supplementary Table 4).

### Association with Psychopathy Symptoms

A total of 14 voxel-wise correlational studies were retrieved, including 12 studies (152 foci, n=1200) and 10 studies (161 foci, n=903) for the Interpersonal/Affective and Chronic Antisocial Lifestyle symptoms, respectively (see Supplementary Table 5). Regional ALE meta-analyses revealed no spatial convergence, suggesting heterogeneous findings between correlational studies. Similarly, using normative network approach no significant effect was identified when using a stringent statistical correction. However, using an unthresholded group-level map, we observed that Interpersonal/Affective symptoms were primarily associated with the Ventral Attention Network (Cohen’s d=.97) and Subcortex (d=.95), whereas the Chronic Antisocial Lifestyle symptoms were rather associated with the Default Mode Network (d=1.11), Subcortex (d=.86), followed by the Somatomotor Network (d=.76) (see Supplementary Figure 3).

## Discussion

The current study aimed to explore for the first time whether the poor replicability of fMRI findings in psychopathy could be enhanced through a network approach. We first conducted a meta-analysis of 23 case-control fMRI studies on psychopathy (534 cases and 593 controls) and found no significant convergence of regional effects. Crucially, however, we demonstrated that the heterogeneous findings across studies are in fact connected to a common functional connectivity network. This network, labeled the Psychopathy Network, is characterized by connectivity hubs including the dorsal anterior cingulate cortex, midcingulate cortex, caudate nucleus, anterior insula, claustrum, lateral prefrontal cortex, thalamus, midbrain, and cerebellum (Lobules VIIIa-b, IX). More importantly, we showed that the Psychopathy Network is highly replicable across studies, reached up to 85.2% overlap in some regions. Employing a multi-method approach to assess its convergent validity, we found strong association between the Psychopathy Network and a lesion network derived from focal brain lesions resulting to antisocial behaviors, as well as with neurotransmission systems (D_1_, 5HTT, DAT) and genetic markers (e.g., OXTR, MAOA), previously implicated in the pathophysiology of psychopathy. Overall, our findings highlight the promise of network mapping in psychopathy, to provide support for and to integrate prior emotion-based ^3–5^, cognitive-based ^6,7^, and neural-based ^8,9,10^ models.

Several neural-based models of psychopathy have been proposed over the past decades, which mainly included the amygdala, vmPFC, ventrolateral PFC, and paralimbic cortical structures ^8–10^. However, discrepancies between the most recent meta-analyses ^15,16^ and broader issue of the lack of replicability across fMRI studies ^46^ have cast doubts on the potential neural markers for psychopathy. Here, we adopted a novel approach to identify associations among heterogeneous brain regions and found that the brain regions initially proposed through theoretical models are replicable and robustly interconnected across studies. For example, Blair ^8^ proposed an integrated emotion systems model where reduced empathic response to the distress of others may result from inadequate pairing between actions that harm others and the aversive consequences of such actions (i.e., aversive stimulus–reinforcement associations). This disruption is posited to be mediated by the amygdala, the vmPFC, the striatum (caudate), dmPFC, and anterior insula ^47^. In our study, we found considerable overlap in brain regions identified in this model (e.g., dmPFC, striatum, anterior insula). Although the vmPFC and the amygdala did not emerge as central functional connectivity hubs, we nevertheless highlighted their significance in the Default Mode Network and Subcortex, which are essential brain networks underpinning socio-affective processes ^48^. Consistent with Blair’s model of psychopathy ^8^, our spatial association analyses on mental functions highlighted the importance of brain regions involved in motivation, value-based decision-making, and socio-affective processes.

Emotion-based models of psychopathy arose from evidence that individuals with psychopathy show reduced autonomic responses to pain and distressing cues ^49^, impaired recognition of fearful emotional expression ^50,51^, and deficits in reinforcement-based decision-making such as reversal learning ^52–54^. However, evidence suggests that the affective deficits observed in psychopathy may be dynamic and context-dependent. Indeed, one core pillar of the cognitive-based models is that individuals with psychopathy show normal affective reactions when instructed to focus their attention on threat-relevant cues, such as when told to increase their emotional response ^55^, take another person’s perspective ^56^ or empathize with other’s emotions ^57^. These cognitive models posit that the affective deficits seen in psychopathy can be explained by their reduced automatic shift of attention to cues that are secondary to their primary focus of attention ^6^ and/or their exaggerated attentional bottleneck, filtering out relevant cues that are in periphery ^7^. Consistent with those modeles, our analysis identified key hubs of the ventral attention and frontoparietal networks, critical for executive functions and attentional shifting (e.g., anterior insula, claustrum, and lateral PFC) ^58^. More importantly, our findings provide a way to reconcile both emotion and cognitive accounts. Indeed, many areas of the Psychopathy Network overlap with core features of autonomic brain activity ^59^ and classical pavlovian fear conditioning ^60^ (i.e., midbrain nuclei, thalamus, pgACC, MCC, and insula/fronto-insular cortex). Given that arousal impacts both cognitive and emotion systems via the ascending arousal network ^61^, these systems are likely intertwined, making them difficult to disentangle. Disruption in connectivity between these distributed sets of regions, rather than localized areas, may thus be a promising avenue to examine the inter-individual variability in psychopathy symptoms Our study significantly contributes to the understanding of psychopathy by presenting a novel approach that integrates emotion-based and cognitive-based models. Indeed, using a multilevel approach to characterize neural correlates of psychopathy, we provide a more comprehensive and complementary understanding of psychopathy and its correlates. However, it should also be noted that psychopathy represents a heterogeneous population, as demonstrated in person-centred (e.g. Primary variant with low anxiety versus Secondary variant with high anxiety) and symptom-centred (e.g., different correlates of interpersonal, affective, lifestyle, and antisocial symptoms) approaches ^1^. Building upon participant-level data, future work should focus on clarifying how clinical heterogeneity relates to the Psychopathy Network, in order to identify novel and more personalized interventions for this disorder.^62^. Lastly, from a developmental perspective, given that psychopathy is increasingly being considered as a neurodevelopmental disorder arising from a complex interplay between genetic and environmental risk factors ^1^, identifying brain networks associated with the development of the disorder will be imperative, both from an aetiological and prevention standpoint. Given that most adults with psychopathy have a history of Conduct Disorder (i.e., a psychiatric disorder characterized by severe antisocial and aggressive behavior in youth), perhaps unsurprisingly the Psychopathy Network we identified appears to share several neural features with the network identified in fMRI studies of Conduct Disorder ^63^ (see also Supplementary Figure 4); this clearly calls for future work using prospective longitudinal studies that ideally start in childhood or prenatally and include multiple levels of analyses ^1^.

### A Search for Biologically-Informed Therapeutic Targets

Despite skepticism among some clinicians regarding the translation of neurobiological findings on psychopathy to clinical practice ^64^, the Psychopathy Network may help identify novel biologically-informed therapeutic targets given its replicability across studies and its associations to specific mental functions, neurotransmission systems and genetic markers. Despite the limited amount of research on biologically-informed treatments for psychopathy, results from psychopharmacological and neuromodulation studies appear promising.

Intranasal oxytocin may be a promising target for the treatment of psychopathy, because of its potential impact on prosocial behavior ^65^ and affiliative emotions (e.g., empathy, attachment) ^66^. For instance, Tully and colleagues ^67^ recently compared 34 violent offenders (19 with psychopathy; 15 without psychopathy) to 24 healthy non-offenders and found that intranasal oxytocin abolished group differences in brain responses to fearful faces in the anterior insula and pMCC, which both appeared as core regions of the Psychopathy Network. Moreover, we showed that OXTR expression across the brain strongly overlapped with regions of the Psychopathy Network (Fig. 4), especially the pMCC, dACC, dmPFC and striatum. Other therapeutic approaches such as neuromodulation could also offer therapeutic avenues, but their application to psychopathy is lacking. For example, Weidacker and colleagues ^68^ applied transcranial direct stimulation to electrode site F4 (10-20 system for EEG electrodes ^69^) during a parametric Go/No-Go task. The authors found that individuals with high cold-heartedness showed better performance during response inhibition, especially under high cognitive load, indicating a potential improvement in attentional control. This site is located near the right lateral PFC identified in our findings, suggesting that its stimulation might alter functional connectivity patterns across a distributed brain network.

In sum, our findings suggest that the Psychopathy Network is strongly associated with molecular (OXTR) and neuromodulation (lateral PFC) targets that could be avenues for future therapeutic interventions. Rather than replacing current psychotherapeutic approaches, these biologically informed treatments may be viewed as complementary. This two-pronged approach to treatment may enhance treatment adherence and engagement (e.g., attentiveness to therapist signals, willingness to establish a working alliance) ^70^.

### Limitations

Despite finding robust evidence that heterogeneous brain regions reported in studies on psychopathy do in fact map onto a common functional connectivity network, we acknowledge several limitations. First, fMRI studies on psychopathy largely differ on a wide range of features including demographics, clinical heterogeneity, MRI protocols and methodological approaches. Despite that we only included case-control studies to preserve homogeneity in analytical approaches between studies it is possible that the Psychopathy Network may have been influenced by other factors. Nevertheless, as previous authors have emphasized, the heterogeneity of psychopathic traits on neurobiological correlates is critical ^71^. We therefore conducted additional analyses by testing whether the Interpersonal/Affective and Chronic Antisocial Lifestyle features of the disorder may be linked to specific connectivity effects. These findings were non-significant, and the interpretation of the unthresholded findings should be approached with caution given the small number of included studies. Second, we used normative resting-state functional connectivity of a large sample of healthy individuals (n=1,000, 50% females) to map the heterogeneous brain regions onto a common network. It is possible that differences in demographics between the included case samples of the included and the normative may have resulted in some variations in network. Future studies using participant-level data should investigate whether matching demographics yield more precise results. Third, we relied on a meta-analytic rather than a mega-analytic approach. International, large-scale collaborative initiatives such as the ENIGMA-Antisocial Behavior Working Group, where individual-level data are shared, may provide a more precise understanding of the neurobiological substrates of Psychopathy that may generalise across populations. Examining network mapping at the individual level may also enable the investigation of the individual differences in the Psychopathy Network which may be used to inform treatments (e.g., molecular, neuromodulation).

## Conclusion

The past two decades have witnessed an intense search for the neural underpinnings of psychopathy, but conflicting findings have cast doubt about the promise of that quest. Here, we show for the first time that heterogeneous peak locations across fMRI studies robustly converge on a common set of cortical and subcortical regions, labelled as the Psychopathy Network. Furthermore, we showed that this network spatially aligns with a lesion network underlying antisocial behaviors and is associated with neurochemical (e.g., dopaminergic, serotonergic systems) and genetic markers (e.g., MAOA, OXTR) previously implicated in the pathophysiology of psychopathy. Taken together, our study highlights the importance of examining the neural correlates of psychopathy from a network perspective, which can be validated using a multilevel approach, encompassing neural, genetic and neurochemical data.

## Methods

### Activation Likelihood Estimation Meta-analysis

A literature search was conducted by reviewing studies included from two recent meta-analyses ^15,16^ and a systematic review ^25^. Studies were included in the current meta-analysis if they: 1) included a sample of adult participants (average >=18 years old); 2) included a case-control analytic approach with groups defined based on a scale assessing psychopathy; 3) included functional magnetic resonance imaging and 4) reported the peak coordinates of the significant group-difference across the whole-brain. Peak coordinates provided in Talairach were then converted onto the MNI standardized space using tal2icbm transformation ^72^. To reduce risk of biases: 1) the impact of multiple experiments per study was handled by concatenating them to form a study-level map ^73^, while the risk of inflating findings through directionality-specific meta-analyses (i.e., conducting meta-analyses for increased and decreased peak activation, separately) was handled by conducting spatial convergence analyses across direction. We conducted an updated coordinate-based meta-analysis of fMRI studies across tasks (see ^15,16^ for previous meta-analyses), using the activation likelihood estimation algorithm (see Supplementary Method). Significance of spatial convergence was established by using a threshold of p<0.001 at voxel-level and FWE-p<0.05 at a cluster-level with 5000 permutations, as recommended ^74,75^. This approach allowed us to identify how many studies report findings that overlap at a voxel-level.

### Normative Network Mapping

We subsequently used a network mapping approach to explore the extent to which heterogeneous peak coordinates across fMRI studies may be linked to a common network ^38,76^. Briefly, a 4-mm sphere was created around each coordinate from each study to create a study-level mask. Then, we computed the normative functional connectivity map of each study-level mask using preprocessed resting-state data of 1,000 healthy subjects (ages 18 to 35 years old, 50% females) of the Brain Genomics Superstruct Project ^77,78^. Information about preprocessing steps is available elsewhere ^78^. We first extracted the time-course of voxels within each of the study-level mask (4mm spheres) and correlated them to the time course of every other voxel in the brain for each of the 1,000 healthy participants, yielding a subject-level study map (1,000 participants x 23 studies). A group-level connectivity map was then computed using a voxel-wise one-sample *t*-test across the 1,000 participants for each study (23 normative maps at a study-level). Replicability at a network-level was established by thresholding each study-level connectivity map at *t* > 5 (as done recently by Stubbs and colleagues^38^ using the same normative sample), binarizing it, and overlapping them. Indeed, others previously found that in some regions, network approaches achieve replicability up to 100% ^38,76^. A subsequent voxel-wise one-sample *t*-test using the unthresholded study-level maps was conducted to generate a Psychopathy Network map that is more consistent than expected by chance through permutation testing ^79^. Main connectivity hubs of this network were identified using cluster-based Threshold-Free Cluster Enhancement (TFCE) and Family-Wise Error corrections (pFWE<0.05).

#### Contribution of Intrinsic Functional Connectivity Networks

Intrinsic functional connectivity networks appear fundamental to characterize signal propagation across the brain. Consequently, we investigated whether our findings may have been driven by particular networks. To do so, we used 8 intrinsic functional connectivity networks, namely the Schaefer-400 parcels 7 Networks ^40^ and a subcortical network ^41^. Effect sizes (Cohen’s *d*) were computed by extracting the averaged t-values across voxels within each intrinsic network and comparing it to the averaged t-values of voxels outside the given network.

### Functional Characterization

#### Brain Lesions Causally Linked to Antisocial Behaviors

Brain lesions are crucial information in understanding causal neurobiological pathways to neurological and psychiatric syndromes. Moreover, early work showed that patients with focal brain lesions manifested a personality disturbance with a similar clinical profile to psychopathy (e.g., “*acquired sociopathy*”, “*pseudopsychopathy*”). Therefore, we examined spatial correlation between the Psychopathy Network and a brain network of lesions that were causally linked to antisocial behaviors ^37^. The latter network was produced by the same approach conducted in our study, using voxels within 17 brain lesions of cases identified through a systematic literature search ^37^. Both Psychopathy Network and Lesion Network were parcellated using brain regions of the 8 intrinsic networks described above, with 7 additional cerebellar regions from the Buckner Atlas ^80^ (Total of 421 regions). Spearman *rho* correlations were then computed on the extracted mean t-values between both sets of brain regions.

#### Mental functions and Neurotransmission systems

We investigated whether the Psychopathy Network may be associated with specific mental functions and neurotransmission systems previously associated with psychopathy. Spatial associations were conducted using JuSpace (version 1.4) ^43^. Psychopathy Network image was first resampled to match the target maps (3mm^3^). The mean values of 421 brain regions (see above) were extracted for the input (Psychopathy-Network [t-values]) and target images (Mental Functions [z-scores] & PET/SPECT [min-max scaled] maps). Partial correlation (*Spearman*’s rank correlation) adjusting for spatial autocorrelation (i.e., local grey matter probabilities) was then performed between the two sets of parcellated maps. Permutation-based p-values (with 5,000 permutations) were computed and further corrected using false discovery rate (FDR) ^81^.

A total of thirteen whole-brain maps of mental functions were used from a data-driven study of more than 1,347 neuroimaging meta-analyses aiming at deriving an ontology of brain circuits spanning various affective, interpersonal and cognitive constructs ^42^. Twenty whole-brain maps representing density of PET/SPECT maps were used ^43,44^. These maps were distributed across 9 neurotransmitter systems including serotonin (i.e., 5-HT_1A_, 5-HT_1B_, 5-HT_2A_, 5-HT_4_, 5-HT_6_, 5-HTT), dopamine (i.e., D_1_, D_2_, DAT), norepinephrine (i.e., NET), Histamine (i.e., H_3_), acetylcholine (i.e., α4β2, M_1_, VAChT), cannabinoid (i.e., CB_1_), opioid (i.e., MOR, KOR), glutamate (i.e., NMDA, mGluR_5_) and GABA (i.e., GABA_A/BZ_).

#### Genetic Markers

We sought to examine whether the Psychopathy Network was significantly associated with sets of genes previously identified across psychiatric disorders using a gene-category enrichment analytical approach. The DisGenNet database^45^ include more than 628 685 gene-disease associations extracted via text mining and expert curation. Genes annotated for 614 disorders and phenotypes from the DisGenNet database was use for the gene-category enrichment analysis. Analyses were conducted using the ABAnnotate toolbox ^82^ which uses brain-wide gene expression patterns obtained from the Allen Human Brain Atlas ^83,84^. The mRNA expression data for 15,633 genes were parcellated into the same 421 brain regions mentioned in the previous section. Null maps of the Psychopathy-Network (5,000) were generated while preserving spatial autocorrelation. Spearman correlations between the Psychopathy Network, the null maps and all the mRNA expression maps were calculated. Positive-sided p-values were then calculated from the comparisons between the “true” category scores with null distribution and were FDR-corrected. To explore the potential role of specific genes, gene-wise spatial associations were then conducted using top genes that characterized disorders associated with the Psychopathy Network.

### Association with Psychopathy Symptoms

Given that the heterogeneity of psychopathy symptoms (i.e., Interpersonal/Affective and Chronic Antisocial Lifestyle) may be linked to specific neural substrates, we conducted an additional literature search using the same criteria as the main search and included studies using a voxelwise correlational approach on psychopathy sub-symptoms. We used the same steps as for the main analyses, which included activation likelihood estimation meta-analysis and normative network mapping for both Interpersonal/Affective and Chronic Antisocial Lifestyle subsymptoms) (see Supplementary Table 5).

## Code Availability

Coordinate-based meta-analysis ALE was conducted using GingerALE version 3.0.2. (http://www.brainmap.org/ale). Network Mapping was conducted using codes from Stubbs and colleagues^38^ (https://github.com/nimlab/NMH_Stubbs2023). Spatial associations with mental functions and neurotransmitters density maps were computed using JuSpace (version 1.4) ^43^ (https://github.com/juryxy/JuSpace). Spatial associations with genetic markers were computed using ABAnnotate toolbox ^82^ (https://github.com/LeonDLotter/ABAnnotate) based on brain-wide gene expression patterns obtained from the Allen Human Brain Atlas ^83,84^ (https://github.com/rmarkello/abagen).

## Data Availability

Peak coordinates of neuroimaging studies included in the meta-analysis can be obtained from the corresponding author on request. Preprocessed resting-state data from the Brain Genomics Superstruct Project is available on the Harvard Dataverse (https://www.neuroinfo.org/gsp).

## Acknowledgements

We would like to thank Dr. Michaël Fox and Dr. Ryan Darby for their generosity in sharing their lesion mask images with us. This study did not receive any specific funding. JRD is holder of a postdoctoral fellowship from the Canadian Institutes of Health Research (MFE-181885). SADB was supported by an Economic and Social Research Council Grant (ES/V003526/1).

