## Supplementary Material for "Mapping the Psychopathic Brain: Divergent Neuroimaging Findings converge onto a Common Brain Network"

Birmingham

**Corresponding authors**

Jules Roger Dugré, PhD

Centre for Human Brain Health, University of Birmingham, School of Psychology, Birmingham B15 2TT

.

&

Stéphane De Brito, PhD;

Centre for Human Brain Health, University of Birmingham, School of Psychology, Birmingham B15 2TT

[**Supplementary Figure 1.** Axial views of the thresholded Psychopathy Network (pFWE<0.05).	10](#_Toc175570032)

[**Supplementary Figure 2.** 98 disorders (DisGeNET) showing gene expression patterns significantly associated with the Psychopathy Network (p<0.001 corrected). Spearman rhos were weighted for the number of genes in each category (i.e., log(number of gene+1))	11](#_Toc175570033)

### **Supplementary Method** Activation Likelihood Estimation

Experiments’ coordinates were used for spatial convergence using the Activation Likelihood Estimate method (GingerALE version 3.0.2, ^1^ http://www.brainmap.org/ale). For each experiment, a 3D gaussian probability distribution was modelled around each coordinate foci, weighted by the number of subjects in the experiment. This method is performed to account for spatial uncertainty due to template and between-subject variance ^2,3^. It also ensures that multiple coordinates from a single experiment does not jointly influence the modeled activation value of a single voxel. The probabilities of all activation foci in an experiment were then combined. Voxel-wise ALE scores arise from the union across all these experiment maps. Consequently, a cluster-level corrected threshold was applied on the voxel-wise image. The size of the supra-threshold clusters was compared against a null distribution of cluster sizes derived from simulation of datasets. We used the following statistical threshold: p<0.001 at voxel-level and FWE-p<0.05 at a cluster-level with 5000 permutations.

### **Supplementary Tables**

| **Supplementary Table 1.** Replicability of the Psychopathy Network Hubs and subregions | | | | | | |
| --- | --- | --- | --- | --- | --- | --- |
| Hub regions | MNI Coordinates | | | Cluster Size (mm^3^) | Cluster-Level Peak Replicability (Max %) | Peak Replicability  (%) |
|  | x | y | z |  |  |  |
| **Positive Connectivity** |  |  |  |  |  |  |
| dmPFC | -20 | 46 | 26 | 35096 | 82.61% | 65.22 |
| dlPFC | -32 | 18 | 40 |  |  | 60.87 |
| dACC | -6 | 20 | 22 |  |  | 60.87 |
| dACC | 6 | 16 | 22 |  |  | 60.87 |
| Anterior Insula/Claustrum | -26 | 18 | -8 | 11976 | 82.61% | 69.56 |
| Caudate | 6 | 10 | 6 |  |  | 60.87 |
| Caudate | -4 | 6 | 6 |  |  | 47.83 |
| Thalamus | -10 | -2 | 0 |  |  | 56.52 |
| Putamen | 32 | 6 | -6 | 2344 | 82.61% | 60.87 |
| vlPFC/AI/Claustrum | 32 | 20 | -12 |  |  | 69.56 |
| Putamen | 24 | 16 | -8 |  |  | 65.22 |
| dlPFC | 32 | 24 | 32 | 6648 | 78.26% | 56.52 |
| dmPFC | 18 | 48 | 28 |  |  | 56.52 |
| SFG | 22 | 38 | 28 |  |  | 52.17 |
| pre-SMA | 14 | 16 | 56 |  |  | 60.87 |
| pMCC | -6 | -18 | 38 | 1520 | 73.91% | 65.22 |
| MCC | 8 | -14 | 36 |  |  | 60.87 |
| **Negative Connectivity** |  |  |  |  |  |  |
| Lobules VIIIb & IX | -18 | -42 | -60 | 47992 | 86.96% | 73.91 |
| Lobule VIIIb | 18 | -42 | -60 |  |  | 65.22 |
| *Note.* dmPFC = dorsomedial Prefrontal Cortex; dACC = dorsal Anterior Cingulate Cortex; vlPFC = ventrolateral Prefrontal Cortex; SFG = Superior Frontal Gyrus; pre-SMA = pre-Supplementary Motor Area; pMCC = posterior MidCingulate Cortex. | | | | | | |

| **Supplementary Table 2.** Spatial Associations between Psychopathy Network and Mental Functions and Neurotransmission maps | | | |
| --- | --- | --- | --- |
| Spatial Maps | z' | p-uncorrected | pFDR |
| Mental Functions | |  |  |
| Social Inference | 0.561 | 0.0002 | 0.0005 |
| Value-Based Decision-Making | 0.556 | 0.0002 | 0.0005 |
| Social Representation | 0.458 | 0.0004 | 0.0007 |
| Motivation | 0.456 | 0.0004 | 0.0007 |
| Physiological Arousal | 0.247 | 0.0002 | 0.0005 |
| Language | 0.233 | 0.0002 | 0.0005 |
| Multiple_Demand | 0.130 | 0.0068 | 0.0104 |
| Spatial Memory | 0.127 | 0.0090 | 0.0117 |
| Auditory Perception | 0.100 | 0.0400 | 0.0473 |
| Face Detection | -0.004 | 0.9306 | 0.9306 |
| Spatial Attention | -0.070 | 0.5151 | 0.5580 |
| Cognitive Control | -0.130 | 0.0072 | 0.0104 |
| Action | -0.211 | 0.0002 | 0.0005 |
| Neurotransmission system | |  |  |
| D_1_ | 0.609 | 0.0002 | 0.0007 |
| 5HTT | 0.597 | 0.0002 | 0.0007 |
| mGLUR_5_ | 0.525 | 0.0006 | 0.0017 |
| DAT | 0.523 | 0.0002 | 0.0007 |
| H_3_ | 0.506 | 0.0008 | 0.0020 |
| VAChT | 0.501 | 0.0002 | 0.0007 |
| MOR | 0.470 | 0.0090 | 0.0180 |
| 5HT_6_ | 0.415 | 0.0002 | 0.0007 |
| NMDA | 0.414 | 0.0012 | 0.0027 |
| KOR | 0.357 | 0.0260 | 0.0433 |
| D_2_ | 0.350 | 0.0002 | 0.0007 |
| α4β2 | 0.326 | 0.0198 | 0.0360 |
| CB_1_ | 0.323 | 0.0316 | 0.0486 |
| 5HT_1A_ | 0.322 | 0.0574 | 0.0703 |
| 5HT_1B_ | 0.296 | 0.0598 | 0.0703 |
| GABA | 0.249 | 0.0598 | 0.0703 |
| 5HT_4_ | 0.229 | 0.0546 | 0.0703 |
| 5HT_2A_ | 0.203 | 0.1900 | 0.2111 |
| M_1_ | 0.127 | 0.3069 | 0.3231 |
| NAT | 0.016 | 0.8324 | 0.8324 |
| *Note.* Spearman Rank (Fisher’s r-to-z transformation). | | | |

| **Supplementary Table 3.** List of Disorders showing significant spatial associations between their categorized gene expression and Psychopathy Network (n=106) | | | | | | |
| --- | --- | --- | --- | --- | --- | --- |
| Disorders | No of Genes | Statistics | | | | |
|  |  | Weighted Correlation  (Fisher's r-to-z) | p-values (uncorrected) | p-values (corr.) | p-values (perm.) | p-values  (perm.+corr) |
| Sexual inhibition | 2 | 0.479 | 1.11E-16 | 6.82E-14 | 0 | 0 |
| Amnestic Disorder | 5 | 0.370 | 8.77E-12 | 2.69E-09 | 0 | 0 |
| Intravenous Drug Abuse | 5 | 0.328 | 2.04E-08 | 1.04E-06 | 0 | 0 |
| Flashing | 2 | 0.325 | 4.76E-07 | 1.22E-05 | 0 | 0 |
| psychiatric hospitalization | 3 | 0.311 | 1.73E-09 | 2.65E-07 | 0 | 0 |
| Emotional Disturbances | 5 | 0.280 | 3.53E-09 | 3.10E-07 | 0 | 0 |
| Psychological addiction | 3 | 0.272 | 7.44E-08 | 2.54E-06 | 0 | 0 |
| Adolescent antisocial behaviour | 2 | 0.264 | 8.02E-07 | 1.63E-05 | 0 | 0 |
| Grandiose delusions | 3 | 0.262 | 2.21E-07 | 6.48E-06 | 0 | 0 |
| Hypervigilance | 3 | 0.260 | 8.06E-07 | 1.63E-05 | 0 | 0 |
| Moderate dementia | 3 | 0.260 | 4.24E-07 | 1.13E-05 | 0 | 0 |
| Cluster C personality disorder | 2 | 0.258 | 1.61E-06 | 2.74E-05 | 0 | 0 |
| Neurotic personality | 2 | 0.258 | 1.61E-06 | 2.74E-05 | 0 | 0 |
| Depression and Suicide | 9 | 0.253 | 5.92E-07 | 1.35E-05 | 0 | 0 |
| Sundowning | 2 | 0.240 | 1.76E-06 | 2.92E-05 | 0 | 0 |
| Delinquent behavior | 16 | 0.238 | 3.35E-09 | 3.10E-07 | 0 | 0 |
| Depressive Syndrome | 34 | 0.236 | 8.21E-09 | 5.60E-07 | 0 | 0 |
| High-functioning autism | 12 | 0.235 | 2.65E-08 | 1.16E-06 | 0 | 0 |
| Severe receptive language delay | 4 | 0.235 | 1.94E-07 | 5.95E-06 | 0 | 0 |
| Polysubstance abuse | 4 | 0.232 | 8.26E-06 | 9.27E-05 | 0 | 0 |
| Melancholia | 34 | 0.229 | 4.75E-08 | 1.82E-06 | 0 | 0 |
| Drug-induced depressive state | 7 | 0.225 | 2.24E-09 | 2.76E-07 | 0 | 0 |
| Gambling, Pathological | 19 | 0.224 | 1.52E-10 | 3.11E-08 | 0 | 0 |
| Psychological Trauma | 7 | 0.223 | 5.79E-07 | 1.35E-05 | 0 | 0 |
| Depression, Neurotic | 31 | 0.222 | 1.41E-08 | 8.65E-07 | 0 | 0 |
| Dysphoric mood | 18 | 0.221 | 2.20E-08 | 1.04E-06 | 0 | 0 |
| Cluster B personality disorder | 3 | 0.220 | 3.95E-06 | 5.51E-05 | 0 | 0 |
| Opioid use disorder, severe | 2 | 0.217 | 8.51E-05 | 5.56E-04 | 0 | 0 |
| Heroin Smoking | 6 | 0.217 | 4.67E-06 | 6.24E-05 | 0 | 0 |
| Hypersexuality state | 6 | 0.215 | 8.22E-07 | 1.63E-05 | 0 | 0 |
| Social Anhedonia | 5 | 0.214 | 9.16E-07 | 1.65E-05 | 0 | 0 |
| Impaired ability to form peer relationships | 2 | 0.214 | 6.39E-05 | 4.41E-04 | 0 | 0 |
| Frigidity | 4 | 0.212 | 1.44E-05 | 1.32E-04 | 0 | 0 |
| Hypoactive Sexual Desire Disorder | 4 | 0.212 | 1.44E-05 | 1.32E-04 | 0 | 0 |
| Orgasmic Disorder | 4 | 0.212 | 1.44E-05 | 1.32E-04 | 0 | 0 |
| Psychosexual Disorders | 4 | 0.212 | 1.44E-05 | 1.32E-04 | 0 | 0 |
| Memory, Short-Term | 8 | 0.209 | 6.89E-07 | 1.51E-05 | 0 | 0 |
| Alcohol problem | 18 | 0.208 | 5.66E-09 | 4.34E-07 | 0 | 0 |
| Fear of heights | 2 | 0.205 | 1.46E-05 | 1.32E-04 | 0 | 0 |
| Addicted to cocaine | 7 | 0.204 | 4.22E-07 | 1.13E-05 | 0 | 0 |
| emotional dysfunction | 7 | 0.201 | 8.75E-07 | 1.65E-05 | 0 | 0 |
| Specific reading disorder | 3 | 0.198 | 3.27E-05 | 2.58E-04 | 0.0002 | 0.0010 |
| Anxiety and fear | 41 | 0.197 | 1.77E-08 | 9.89E-07 | 0 | 0 |
| Narcissism | 7 | 0.196 | 6.00E-06 | 7.55E-05 | 0 | 0 |
| Autotomy | 2 | 0.194 | 4.15E-05 | 3.04E-04 | 0 | 0 |
| Ecstasy related disorders | 10 | 0.191 | 7.08E-06 | 8.36E-05 | 0 | 0 |
| Completed Suicide | 26 | 0.188 | 6.34E-08 | 2.29E-06 | 0 | 0 |
| Compulsive sexual behaviour | 2 | 0.188 | 5.61E-05 | 4.01E-04 | 0 | 0 |
| Outbursts | 2 | 0.186 | 1.96E-05 | 1.67E-04 | 0 | 0 |
| Psychological pseudocyesis | 8 | 0.183 | 1.31E-05 | 1.32E-04 | 0 | 0 |
| Stereotypical body rocking | 3 | 0.182 | 1.13E-04 | 6.97E-04 | 0 | 0 |
| Aggressive outburst | 2 | 0.181 | 1.75E-05 | 1.53E-04 | 0 | 0 |
| Mania acute | 2 | 0.181 | 3.92E-05 | 2.90E-04 | 0 | 0 |
| Catatonia | 12 | 0.178 | 1.42E-05 | 1.32E-04 | 0 | 0 |
| Impulsive character (finding) | 32 | 0.178 | 1.00E-07 | 3.24E-06 | 0 | 0 |
| Dysthymic Disorder | 8 | 0.174 | 2.54E-06 | 3.90E-05 | 0 | 0 |
| Food Addiction | 7 | 0.172 | 3.77E-05 | 2.86E-04 | 0 | 0 |
| Antisocial behavior | 34 | 0.172 | 2.31E-06 | 3.64E-05 | 0 | 0 |
| Prolonged grief disorder | 3 | 0.168 | 7.38E-06 | 8.55E-05 | 0 | 0 |
| Paranoid Schizophrenia | 34 | 0.167 | 2.82E-08 | 1.16E-06 | 0 | 0 |
| Premenstrual Dysphoric Disorder | 8 | 0.166 | 2.07E-05 | 1.74E-04 | 0 | 0 |
| Algophobia | 6 | 0.166 | 3.74E-06 | 5.44E-05 | 0 | 0 |
| Spastic | 13 | 0.165 | 6.34E-05 | 4.41E-04 | 0 | 0 |
| Atypical autism | 3 | 0.165 | 9.89E-05 | 6.39E-04 | 0.0002 | 0.0010 |
| Mixed dementia | 7 | 0.164 | 1.79E-05 | 1.55E-04 | 0 | 0 |
| Psychoses, Drug | 21 | 0.161 | 5.98E-06 | 7.55E-05 | 0 | 0 |
| ADHD, combined type | 5 | 0.159 | 3.62E-05 | 2.78E-04 | 0.0002 | 0.0010 |
| Other and unspecified reactive psychosis | 6 | 0.157 | 4.98E-05 | 3.60E-04 | 0 | 0 |
| Sexual Arousal Disorder | 5 | 0.156 | 1.38E-04 | 8.23E-04 | 0.0002 | 0.0010 |
| Endogenous depression | 38 | 0.154 | 6.89E-06 | 8.29E-05 | 0 | 0 |
| Mild dementia | 10 | 0.154 | 2.34E-05 | 1.94E-04 | 0 | 0 |
| Processing speed | 4 | 0.154 | 1.32E-04 | 7.92E-04 | 0.0002 | 0.0010 |
| Neurocognitive Disorders | 56 | 0.151 | 8.95E-07 | 1.65E-05 | 0 | 0 |
| Severe dementia | 12 | 0.149 | 1.05E-05 | 1.11E-04 | 0 | 0 |
| parental alcoholism | 4 | 0.148 | 1.04E-04 | 6.59E-04 | 0.0002 | 0.0010 |
| Anxiety neurosis (finding) | 48 | 0.146 | 5.75E-07 | 1.35E-05 | 0 | 0 |
| Violence | 46 | 0.144 | 4.26E-06 | 5.82E-05 | 0 | 0 |
| Postoperative delirium | 33 | 0.140 | 1.48E-04 | 8.74E-04 | 0 | 0 |
| Minimal Brain Dysfunction | 18 | 0.140 | 7.40E-05 | 4.94E-04 | 0 | 0 |
| Disturbance in mood | 18 | 0.139 | 1.04E-05 | 1.11E-04 | 0 | 0 |
| Sexual Dysfunction | 24 | 0.138 | 8.30E-06 | 9.27E-05 | 0 | 0 |
| Repetitive compulsive behavior | 6 | 0.135 | 7.10E-05 | 4.79E-04 | 0.0002 | 0.0010 |
| heroin abuse | 14 | 0.135 | 1.08E-04 | 6.76E-04 | 0 | 0 |
| Anxiety States, Neurotic | 32 | 0.134 | 1.33E-05 | 1.32E-04 | 0 | 0 |
| Recurrent depressive disorder | 16 | 0.134 | 1.36E-05 | 1.32E-04 | 0 | 0 |
| Anxiety state | 23 | 0.134 | 3.00E-05 | 2.40E-04 | 0 | 0 |
| Opioid withdrawal | 14 | 0.132 | 8.25E-05 | 5.45E-04 | 0 | 0 |
| Panic Disorder | 112 | 0.131 | 2.98E-06 | 4.46E-05 | 0 | 0 |
| manic symptom | 14 | 0.130 | 1.62E-04 | 9.40E-04 | 0.0006 | 0.0028 |
| Psychoses, Substance-Induced | 15 | 0.129 | 1.03E-04 | 6.59E-04 | 0 | 0 |
| Catalepsy | 31 | 0.128 | 6.50E-05 | 4.44E-04 | 0 | 0 |
| Aphasia, Progressive | 8 | 0.128 | 6.70E-06 | 8.23E-05 | 0 | 0 |
| Bulimia Nervosa | 47 | 0.127 | 3.36E-05 | 2.61E-04 | 0 | 0 |
| Anorexia Nervosa | 137 | 0.127 | 1.95E-06 | 3.15E-05 | 0 | 0 |
| Obsessive-Compulsive Disorder | 124 | 0.126 | 6.03E-06 | 7.55E-05 | 0 | 0 |
| Cocaine-Related Disorders | 91 | 0.125 | 6.07E-05 | 4.28E-04 | 0 | 0 |
| Delirium, Dementia, Amnestic Disorders | 60 | 0.125 | 1.56E-05 | 1.38E-04 | 0 | 0 |
| Conduct Disorder | 25 | 0.120 | 1.18E-05 | 1.22E-04 | 0 | 0 |
| Depressive episode, unspecified | 17 | 0.119 | 2.89E-05 | 2.33E-04 | 0 | 0 |
| Depressed bipolar I disorder | 33 | 0.115 | 1.04E-05 | 1.11E-04 | 0 | 0 |
| Antisocial Personality Disorder | 44 | 0.106 | 3.87E-05 | 2.89E-04 | 0 | 0 |
| Memory dysfunction | 60 | 0.104 | 3.81E-06 | 5.44E-05 | 0 | 0 |
| Executive dysfunction | 27 | 0.103 | 1.13E-04 | 6.97E-04 | 0 | 0 |
| Aloof | 61 | 0.101 | 1.27E-04 | 7.75E-04 | 0.0002 | 0.0010 |
| Apraxia, Developmental Verbal | 53 | 0.100 | 1.55E-04 | 9.09E-04 | 0 | 0 |
| Social Anxiety | 44 | 0.095 | 2.41E-05 | 1.97E-04 | 0 | 0 |
| Note.Threshold was set at p<0.001 corrected which identified 106 disorders. | | | | | | |

| **Supplementary Table 4.** Associations between brain expression of identified genes and Psychopathy Network (>10% occurrence across the 106 identified disorders) | | | |
| --- | --- | --- | --- |
| Genes | Frequency | Spearman Rho | p-value  (uncorrected) |
| BDNF | 37.74% | 0.190 | 2.95E-04 |
| COMT | 30.19% | 0.188 | 3.49E-04 |
| DRD2 | 29.25% | 0.085 | 1.09E-01 |
| MAOA | 24.53% | 0.213 | 4.72E-05 |
| SLC6A3 | 20.75% | -0.003 | 9.59E-01 |
| APOE | 19.81% | 0.210 | 5.90E-05 |
| HTR1A | 17.92% | 0.230 | 1.07E-05 |
| CNR1 | 16.98% | 0.271 | 1.78E-07 |
| DRD4 | 16.98% | 0.019 | 7.14E-01 |
| OPRM1 | 14.15% | 0.218 | 3.06E-05 |
| OXTR | 14.15% | 0.230 | 1.08E-05 |
| CRH | 12.26% | 0.079 | 1.34E-01 |
| HTR2A | 12.26% | -0.117 | 2.71E-02 |
| TNF | 12.26% | -0.253 | 1.25E-06 |
| ELK3 | 10.38% | 0.085 | 1.10E-01 |
| GABRA2 | 10.38% | 0.253 | 1.24E-06 |
| NPY | 10.38% | 0.167 | 1.51E-03 |
| NR3C1 | 10.38% | -0.310 | 1.86E-09 |
| Note. | | | |

| **Supplementary Table 5.** Characteristics of the included voxelwise correlational studies (k=14) | | | | | | | | |
| --- | --- | --- | --- | --- | --- | --- | --- | --- |
| First Author, Year | Sample Characteristics | | | | Psychopathic Traits | | | Task |
|  | Age | N | Males  (%) | Setting | Measures | Factor 1 | Factor 2 |  |
| Anderson, 2017 ^4^ | 32.9 | 120 | 100.0% | Correctional | PCL-R | X | - | Implicit Emotion Processing |
| Anderson, 2018 ^5^ | 37.1 | 168 | 100.0% | Correctional | PCL-R | X | X | Auditory Oddball Task |
| Caldwell, 2015 ^6^ | 33.6 | 87 | 100.0% | Correctional | PCL-R | X | - | Implicit Morality Processing |
| Contreras-Rodriguez, 2014 ^7^ | 39.8 | 22 | 100.0% | Correctional | PCL-R | X | X | Emotional face-matching task |
| Cope, 2014 ^8^ | 34.0 | 137 | 32.1% | Correctional | PCL-R | X | X | Cue-Elicited Craving Task |
| Decety, 2013 ^9^ | 18-50 | 80 | 100.0% | Correctional | PCL-R | X | X | Painful Situations and Expressions |
| Decety, 2015 ^10^ | 19-54 | 155 | 100.0% | Correctional | PCL-R | X | X | Morally-Laden Scenarios |
| Deming, 2018 ^11^ | 18-55 | 57 | 100.0% | Correctional | PCL-R | - | X | Trait judgment task |
| Deming, 2020 ^12^ | 32.9 | 94 | 100.0% | Correctional | PCL-R | - | X | Affective Perspective-Taking Task |
| Denomme, 2018 ^13^ | 38.3 | NA | NA | Correctional | PCL-R | X | - | Cue-Elicited Craving Task |
| Harenski, 2014 ^14^ | 33.2 | 157 | 0.0% | Correctional | PCL-R | X | X | Implicit Morality Processing |
| Sadeh, 2013 ^15^ | 33.7 | 49 | 38.8% | Community | NEO-FFI | X | X | Emotional Stroop |
| Yoder, 2015 ^16^ | 31.2 | 94 | 100.0% | Correctional | PCL-R | X | X | Morality Processing |
| Yoder, 2021^17^ | 35.0 | 107 | 0.0% | Correctional | PCL-R | X | X | Moral Interactions |
| Note. NA = Not Available | | | | | | | | |

### **Supplementary Figures**

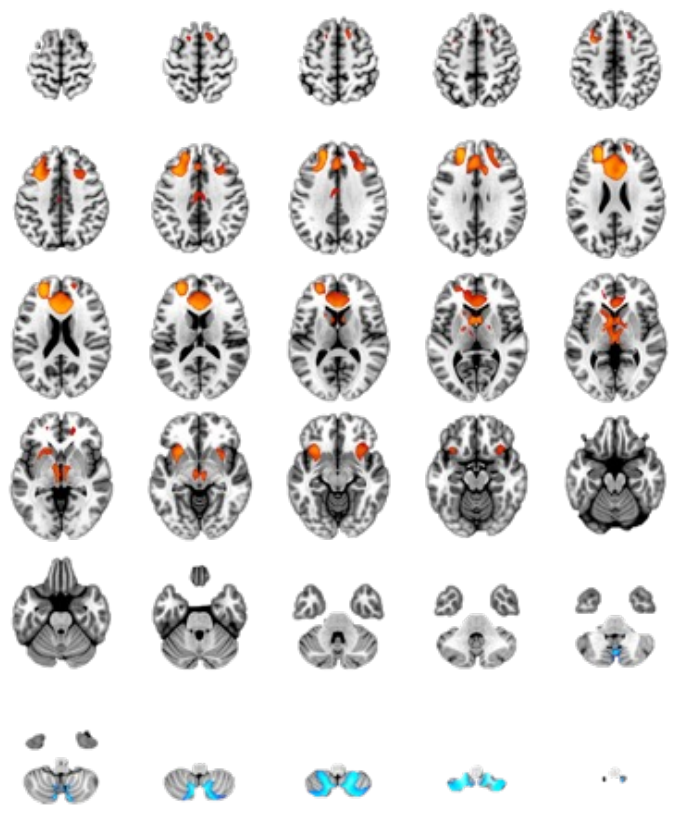

**Supplementary Figure 1.** Axial views of the thresholded Psychopathy Network (pFWE<0.05).

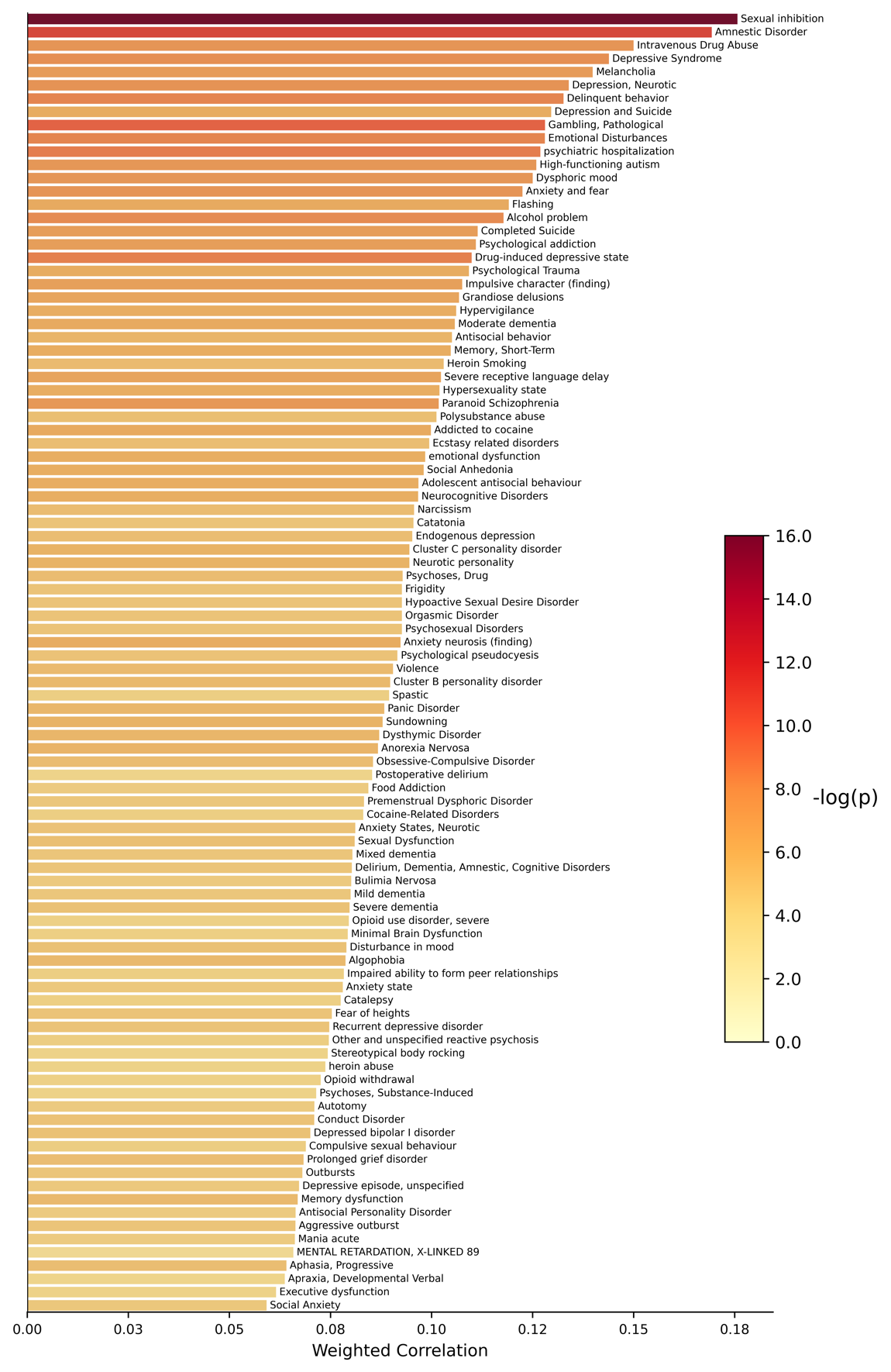

#### **Supplementary Figure 2.** 98 disorders (DisGeNET) showing gene expression patterns significantly associated with the Psychopathy Network (p<0.001 corrected). Spearman rhos were weighted for the number of genes in each category (i.e., log(number of gene+1))

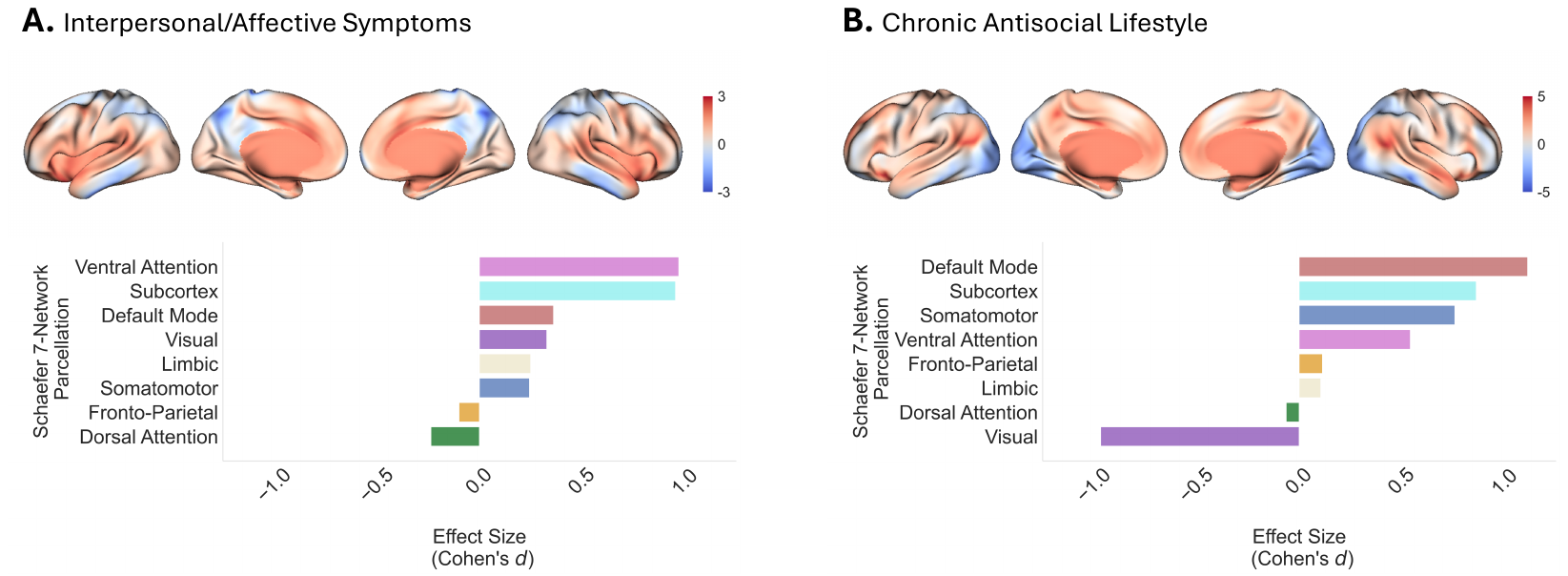

**Supplementary Figure 3.** Unthresholded voxelwise functional connectivity maps of the Interpersonal/Affective and Chronic Antisocial Lifestyle symptoms of Psychopathy. Figures also display the contribution of 8 intrinsic networks including Schaefer-400 parcels 7-Network and subcortex.

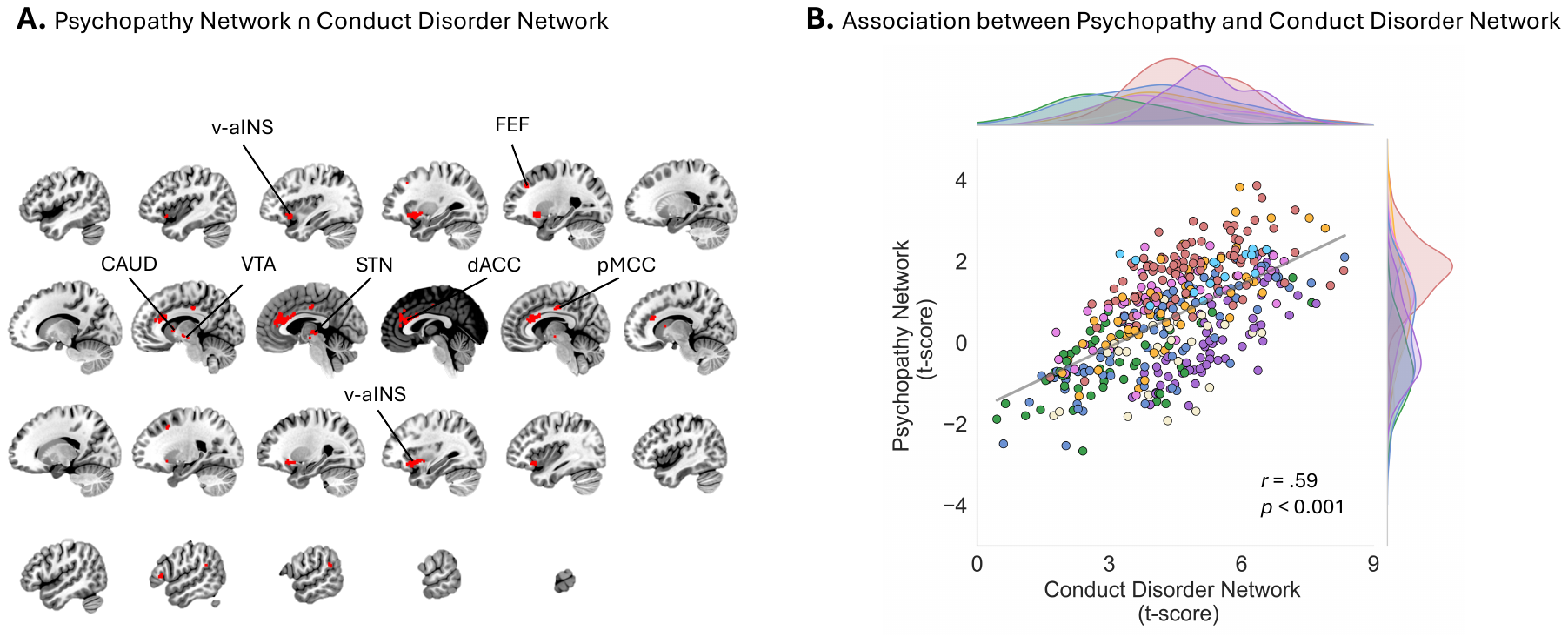

**Supplementary Figure 4.** Spatial overlap between Psychopathy Network and Conduct Disorder (Dugré, Potvin, 2024). **A.** Spatial overlap of brain regions showing high replicability across studies of for both Psychopathy and Conduct Disorder (greater than 60% replicability). v-aINS = ventral-anterior Insula; FEF = Frontal Eye Fields; CAUD = Caudate Nucleus; VTA = Ventral Tegmental Area; STN = Subthalamic Nuclei; dACC = dorsal Anterior Cingulat Cortex; pMCC = posterior MidCingulate Cortex; **B.** Spatial correlation between unthresholded t-maps of Psychopathy Network and Conduct Disorder Network (Spearman rho = .59, p<0.001). t-maps were parcellated using 421 regions described in the main manuscript.
